# Interrelations Between Dopaminergic-, GABAergic- and Glutamatergic Neurotransmitters in Antipsychotic-Naïve Psychosis Patients and the Association to Initial Treatment Response

**DOI:** 10.1101/2025.02.18.25322467

**Authors:** Kirsten Borup Bojesen, Karen S. Ambrosen, Anne Korning Sigvard, Mette Ødegaard Nielsen, Albert Gjedde, Yoshitaka Kumakura, Lars Thorbjørn Jensen, Dan Fuglø, Bjørn H. Ebdrup, Egill Rostrup, Birte Yding Glenthøj

## Abstract

Preclinical evidence points to disturbances in neural networks in psychosis involving interrelations between dopaminergic-, GABAergic- and glutamatergic neurotransmitter systems. In support, we have previously shown that aberrant interrelations between these neurotransmitters, in contrast to individual transmitter systems, can separate antipsychotic-naïve first-episode psychotic patients (AN-FEP) from healthy controls (HC). Here, we characterized neurotransmitter interrelations, examined their association with treatment response, and explored the effect of treatment on the interrelations.

Sixty participants (29 AN-FEP and 31 HC) underwent dynamic [18F]-DOPA PET with arterial blood sampling using a four-parameter model to measure dopamine synthesis (DS) (k_3_) in nucleus accumbens (NAcc) and magnetic resonance spectroscopy (MRS) to estimate levels of glutamate (Glu) in anterior cingulate cortex (ACC) and thalamus, and gamma-aminobutyric-acid (GABA) in ACC. A subgroup of the patients was re-scanned after six weeks antipsychotic monotherapy with aripiprazole. Psychopathology was assessed at both visits. Multiple linear regression models and linear mixed models were used to analyze data.

We found a negative association between k_3_ (dependent variable) and GABA in HC (β=-0.15, p=0.03) and a positive association in patients (β=0.15, p=0.04). The aberrant relationship between k_3_ and GABA was driven by the group-GABA interaction (p=0.002) and related to treatment response (p=0.02). No significant group interactions were found for the interrelations between k_3_ and Glu, but a positive association was found between k_3_ and Glu in thalamus (p=0.04) in both groups and the association decreased after treatment in AN-FEP (p=0.01).

The data show that DS in NAcc and GABA levels in ACC are inversely interrelated in AN-FEP, and that the degree of abnormality predicts treatment effect. Moreover, antipsychotic treatment alters the relationship between dopaminergic activity in NAcc and Glu levels in thalamus. The findings suggest that combined instead of single neurotransmitter disturbances should be considered when novel therapeutics are developed for schizophrenia.

## Introduction

Aberrations in the interactions and connections (hereafter interrelations) between dopaminergic-, gamma-aminobutyric acid (GABA)- and glutamatergic neurotransmitter systems in the cortico-striato-thalamo-cortical macro-circuits have for decades been suggested to be involved in the development of psychosis as well as the effect of treatment (1–6). While these interrelations are well described in preclinical literature, they are sparsely studied in clinical studies that primarily have reported on abnormalities of single neurotransmitters. Overall, the findings do not support altered post-or presynaptic dopamine activities in antipsychotic-naïve first-episode psychotic patients (AN-FEP) (7–11), whereas increased activity is seen in medicated or chronic patients (12–15). For GABA levels in anterior cingulate cortex and nearby areas (ACC hereinafter), levels appear decreased at illness onset but seem to normalize in the chronic stage (16–19).

Glutamate (Glu) levels in ACC seem to be decreased in first-episode patients, whereas subcortical Glu levels in striatum and thalamus appear to be increased (17–22). Combined, these results suggest an abnormal relationship between neurotransmitters in the cortico-striato-thalamo-cortical macro-circuits in patients with psychosis, presumably due to decreased prefrontal glutamatergic and GABAergic regulation of striatal and thalamic activity. Still, clinical studies of combined neurotransmitter disturbances are needed before conclusions can be drawn. So far, only two studies investigated the combined relationships between striatal activity and ACC Glu levels in FEP (11) and healthy control subjects (HC) (23), respectively. The first study reported a negative association between ACC Glu levels and striatal dopamine synthesis capacity (DSC) in first-episode patients that was not present in HC (11), whereas the other study found a negative association between prefrontal Glu levels and striatal DSC in HC (23). To date, only one study has reported on ACC GABA levels in first-episode psychosis patients and striatal perfusion as a proxy-measure of dopaminergic activity (16). In that study, we found that higher levels of the inhibitory neurotransmitter GABA were related to lower striatal perfusion in both AN-FEP and HC (16). However, more direct measures are needed to reveal abnormalities between prefrontal GABA levels and striatal dopaminergic activity in AN-FEP, and to our knowledge, such studies have not been published this far.

To address the impact of multiple neurotransmitter measures on the schizophrenia pathology, we recently tested if the combination of presynaptic striatal dopamine activity, levels of Glu and GABA in ACC, and levels of Glu in left thalamus could separate AN-FEP from matched HC (24). In line with the literature, patient status could not be predicted based on individual neurotransmitters, whereas a combination of dopamine activity in nucleus accumbens (NAcc), ACC GABA levels, and thalamic Glu levels separated AN-FEP from HC with an accuracy of 83.7% where, especially, the interaction between striatal dopaminergic activity and prefrontal GABA levels contributed to patient identification (24). Adding thalamic Glu levels increased the accuracy, although thalamic Glu levels in itself did not predict patient status (24). These findings support that combined measures of dopaminergic activity in striatum, GABA levels in ACC, and Glu levels in thalamus are crucial to the schizophrenia pathophysiology, but it still needs to be investigated how the relationship between dopaminergic activity in NAcc and prefrontal GABA and thalamic Glu levels differs between AN-FEP and HC.

Treatment response to antipsychotics may also depend on combined neurotransmitter disturbances, but up till now, studies have only investigated associations with single neurotransmitter levels. Antipsychotics dampen dopamine activity via dopamine D_2_ receptors (25), and treatment effect has been related to the availability of frontal and striatal D_2_ receptors in AN-FEP (8–10) as well as to presynaptic striatal dopamine activity in FEP (7, 11). In the largest longitudinal positron emission tomography (PET) study of AN-FEP to date, we used arterial blood sampling and a four-parameter (4P) model to study striatal dopamine synthesis (DS: the decarboxylation rate of [18F]-DOPA to [18F]-dopamine = k_3_) and DSC (K ^4p^), and, for comparison, we also assessed Ki^cer^ values with the commonly used tissue reference (TR) method (7). Using the 4P model, we found highly significant associations between k_3_ estimates in nucleus accumbens (NAcc) and psychotic symptoms at baseline as well as between k_3_ and the effect of six weeks of treatment with the partial D_2_ receptor agonist aripiprazole, whereas no associations were found with the TR-method (7). This points to k_3_ estimates as key markers of psychopathology as well as treatment response in psychosis.

Previous data from an overlapping group of AN-FEP subjects likewise showed that increased levels of Glu in thalamus and decreased levels of GABA in ACC were related to treatment response (19). Other studies reported glutamatergic abnormalities in ACC in first-episode patients with poor treatment response (26, 27), supporting a link between glutamatergic abnormalities and treatment effect of first-line treatment, although the affected region and direction of abnormality differs. However, it has never been possible to separate AN-FEP from HC subjects based on single transmitters, but only by using combined measures of dopaminergic activity in striatum, GABA levels in ACC, and Glu levels in thalamus, where especially the interactions between dopaminergic activity in NAcc and GABA levels in ACC was important (24). An aberrant interrelation between striatal dopaminergic activity and prefrontal GABA levels may therefore be relevant for treatment response as well. Further, a recent study has suggested that antipsychotic treatment alters the neurotransmitter relationship between striatal DSC and Glu levels in ACC (28). However, no previous study has investigated whether treatment alters the interrelations between presynaptic striatal dopamine activity and GABA levels in ACC or Glu levels in thalamus.

Our primary aim was to study how the interrelations between striatal DS and cortical GABA levels differ between strict AN-FEP and matched HC. We further explored if combined measures of GABA and Glu levels in ACC and Glu levels in thalamus characterized striatal DS in AN-FEP more accurately than GABA levels alone. Our secondary aim was to investigate if an abnormal relationship between striatal DS and GABA levels in ACC has an impact on treatment effect with a partial dopamine agonist aripiprazole. In explorative analyses we additionally investigated if the interrelations between striatal DS and GABA as well as glutamate levels change after treatment.

We hypothesized inverse interrelations between DS (k_3_) in NAcc and GABA levels in ACC at baseline associated with the subsequent effect of treatment on psychotic symptoms. In previous studies of individual neurotransmitter levels, neither dopamine activity in NAcc (7) nor Glu levels in ACC (19) changed following treatment with aripiprazole whereas this was the case for thalamic Glu levels (19); accordingly, we also expect changes in the relation between k_3_ and thalamic Glu following treatment.

## Methods and Materials

### Participants

Participants were part of a large multimodal cohort study (PECANS II) approved by the National Committee on Biomedical Research Ethics (H-3-2013-149) previously described in detail (7, 19).

Patients were referred from Mental Health Centers in the Capital Region of Denmark. Inclusion criteria were: lifetime antipsychotic-naïve; lifetime naïve to central nervous system stimulants, 18-45 years of age, legally competent, and fulfilling the diagnostic criteria for schizophrenia, schizoaffective disorder, or non-organic psychosis according to the International Classification of Diseases, 10^th^ revision (ICD-10) as evaluated by certified interviewers with the ‘Schedules for Clinical Assessment in Neuropsychiatry’ (SCAN)

(29). We assessed psychopathology with the Positive and Negative Syndrome Scale (PANSS)(30) at baseline and after six weeks of treatment with flexible doses of a partial D_2_ agonist (aripiprazole). Treatment effect was estimated as change of PANSS positive scores, denoted ΔPANSS, from baseline to six weeks’ follow-up (PANSS positive baseline - PANSS positive follow-up). Hence, a negative ΔPANSS indicates an improvement of positive symptom severity. Medication compliance was assessed by serum concentrations.

We recruited HC matched on age, sex, and parental educational level through online advertisement (www.forsøgsperson.dk) with the following exclusion criteria: not being in ultra-high-risk for psychosis according to the Comprehensive Assessment of At-Risk Mental States (31), no previous mental health issues and not having a first degree relative with mental illness (lifetime).

Patients were excluded in case of involuntarily admission or treatment or if they had been treated with antidepressant during the last 30 days or were in need of antidepressants between baseline and six-week follow-up examinations. All participants were excluded if they had experienced a head injury with more than 5 minutes of unconsciousness, had metallic implants (not compatible with MRI), were pregnant, had a severe physical illness, or previous substance abuse. Prescribed benzodiazepines were tolerated in patients before initiation of antipsychotic treatment after examinations, although not 12 hours before magnetic resonance imaging (MRI) or PET-imaging. Substance use was assessed through self-report and verified by a urine drug test (Rapid Response, Jepsen HealthCare, Tune, Denmark) before PET and magnetic resonance spectroscopy (MRS).

### [18F]-DOPA PET

DS was assessed with [18F]-DOPA PET imaging with integrated PET-CT (Siemens Biograph m CT64 from 2013). We administered carbidopa 150 mg and entacapone 400 mg orally one hour before PET to minimize [18F]-DOPA metabolic degradation before passage through the blood brain barrier. A low-dose CT-scan was performed before each PET-session to enable attenuation correction. A detailed description of the methodology is provided elsewhere (7).

### Magnetic Resonance Imaging and Magnetic Resonance Spectroscopy

Proton magnetic resonance spectroscopy (1H-MRS) and T1 weighted structural MRI were performed on a 3.0 Tesla scanner (Achieva, Philips Healthcare, Eindhoven, NL) with a 32-channel head coil (Invivo, Orlando, Lorida, USA) as previously described (32). A T1 weighted structural scan (TR: 10ms; TE: 4.6ms; flip angle: 8°; voxel size: 0.79*0.79*0.80mm^3^) was obtained for co-registration and anatomical reference of the PET images as well as segmentation of gray- and white matter in the spectroscopic voxels. We used FreeSurfer (13,14) version 5.3.0 (33, 34) for individual segmentation in the PET analyses, where nucleus accumbens (NAcc) was the primary region of interest (ROI) based on previous data (7).

Point-resolved spectroscopy (PRESS) was used to estimate levels of Glu (TR 3000ms, TE 30ms, 128 averages with MOIST water-suppression, 7 min pr. Scan). Spectra were obtained in a 2.0×2.0×2.0cm^3^ voxel in dorsal ACC and a 2.0×1.5×2.0cm^3^ voxel in left thalamus (Supplementary Figure S1 B and C) simultaneously with an inbuilt unsuppressed water reference scan. Glu levels were estimated by fitting spectra in the range of 0.2-4.0ppm using LCModel version 6.3-1L (s-provencher.com/lcmodel.shtml) (35).

Last, Mescher–Garwood point-resolved spectroscopy sequence (MEGAPRESS) was used for acquisitions of GABA levels (TE=68ms; TR=2000ms, 14ms editing pulses applied at 1.9 and 7.5ppm, 320 averages, MOIST water suppression, and interleaved unsuppressed water reference) (36) in a 3.0×3.0×3.0cm^3^ voxel placed in dorsal ACC (Supplementary Figure S1 A). Gannet version 3.1 was used to fit GABA levels quantified as water-scaled values in the spectral range between 2.79 and 3.55ppm (37).

All metabolite levels were calculated in institutional units by correcting for partial volume cerebrospinal fluid as previously described (18). Minimum reporting standards for MRS (38) including quality data are provided in Supplementary Tables S1, S2, S3, and S4. In ACC, there were small but significant differences in FWHM for PRESS acquisitions, and fit error as well as FWHM for MEGAPRESS acquisitions at baseline (Supplementary Table S3 and S4) but no other significant group differences in data quality parameters.

### Statistical analysis

Multiple linear regression using Ordinary Least Squares and linear mixed effects model analyses were performed in Python version 3.12 as implemented in the statsmodels library version 0.14.1. Independent variables were standardized by subtracting the mean and dividing by the standard deviation prior to the analyses to minimize collinearity. Group differences in demographic data were analyzed using Mann-Whitney U test, Fischeŕs exact test, and Chi-square tests as appropriate in SAS version 8.4.

### Group differences in neurotransmitter interrelations

Separate multiple linear regression analyses were applied to test the primary hypothesis that the interrelations between DS in NAcc (k_3_) and ACC GABA levels differed in AN-FEP and matched HC. First, the relationship between k_3_ in NAcc as dependent variable (y) and GABA in ACC, sex, and GABA*group was tested (Eq. 1).

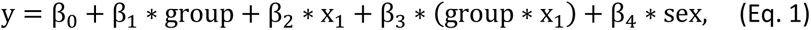

where β_0_is the intercept, x_1_ is GABA in ACC, and β_1_ − β_4_ are the coefficients of the independent variables. In case of a significant GABA*group interaction, post hoc tests were performed for AN-FEP and HC separately using general linear models.

Second, we examined the additive effect of combinations of neurotransmitters by fitting models including the independent variables group, GABA in ACC, group-GABA interaction, and either Glu in ACC or Glu in thalamus in the same model (Eq. 2).

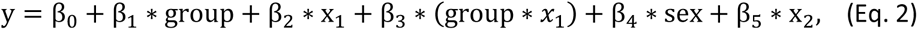

where x_1_ is GABA in ACC and x_2_ are either Glu in ACC, or Glu in thalamus.

In exploratory analyses, we tested the relationship between k_3_ in NAcc and either Glu in thalamus or Glu in ACC in a similar model as Eq. 1.

### The relation between treatment response and neurotransmitter interrelations

Separate multiple linear regression analyses tested the second hypothesis that an abnormal relationship between striatal DS and GABA levels in ACC was associated with treatment response. First, the relationship between ΔPANSS positive as the dependent variable (y), and k_3_, GABA levels, and the k_3_*GABA interaction as independent variables (*x*_1_and *x*_2_, and *x*_1_ ∗ *x*_2_) was examined with adjustment for the covariate sex that previously has been shown to influence the load of positive symptoms (9) (Eq. 3).

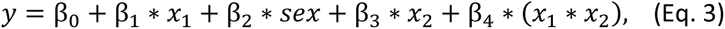

where β_0_is the intercept, and β_1_ − β_4_ are the coefficients of the independent variables. In explorative analyses, similar models were tested by exchanging GABA levels (*x*_2_) with Glu levels in either thalamus or ACC.

### Changes in neurotransmitter interrelations after treatment

In explorative analyses in the patient group, separate linear mixed effects analyses were performed to examine the effects of treatment on the neurotransmitters and the relationship between them. First, we explored whether the association between k_3_ and either GABA in ACC, Glu in thalamus, or Glu in ACC changed after treatment. In these analyses, k_3_ was the dependent variable (y). The neurotransmitter of interest (x_1_), visit (baseline vs follow-up), and the neurotransmitter*visit interaction were included as fixed effects, with a random participant-level effect (Eq. 4). The baseline visit was used as the reference.

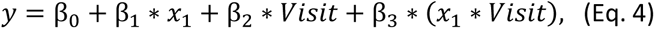

where β_0_is the intercept, and β_1_ − β_3_ are the coefficients of the independent variables. Further, the effect of treatment on each neurotransmitter individually was explored. In these models the neurotransmitter of interest was the dependent variable, visit was a fixed effect, with a random participant-level effect (i.e., x_1_=0 in Eq. 4).

## Results

A total of 62 participants (31 patients and 31 HCs) were included. Of these, two patients were excluded due to a positive screening for benzodiazepines on neuroimaging days, leaving a study population of 29 patients and 31 HCs as shown in Figure 1 together with a study flow-chart illustrating usable PET and MRS data.

**Figure 1:**
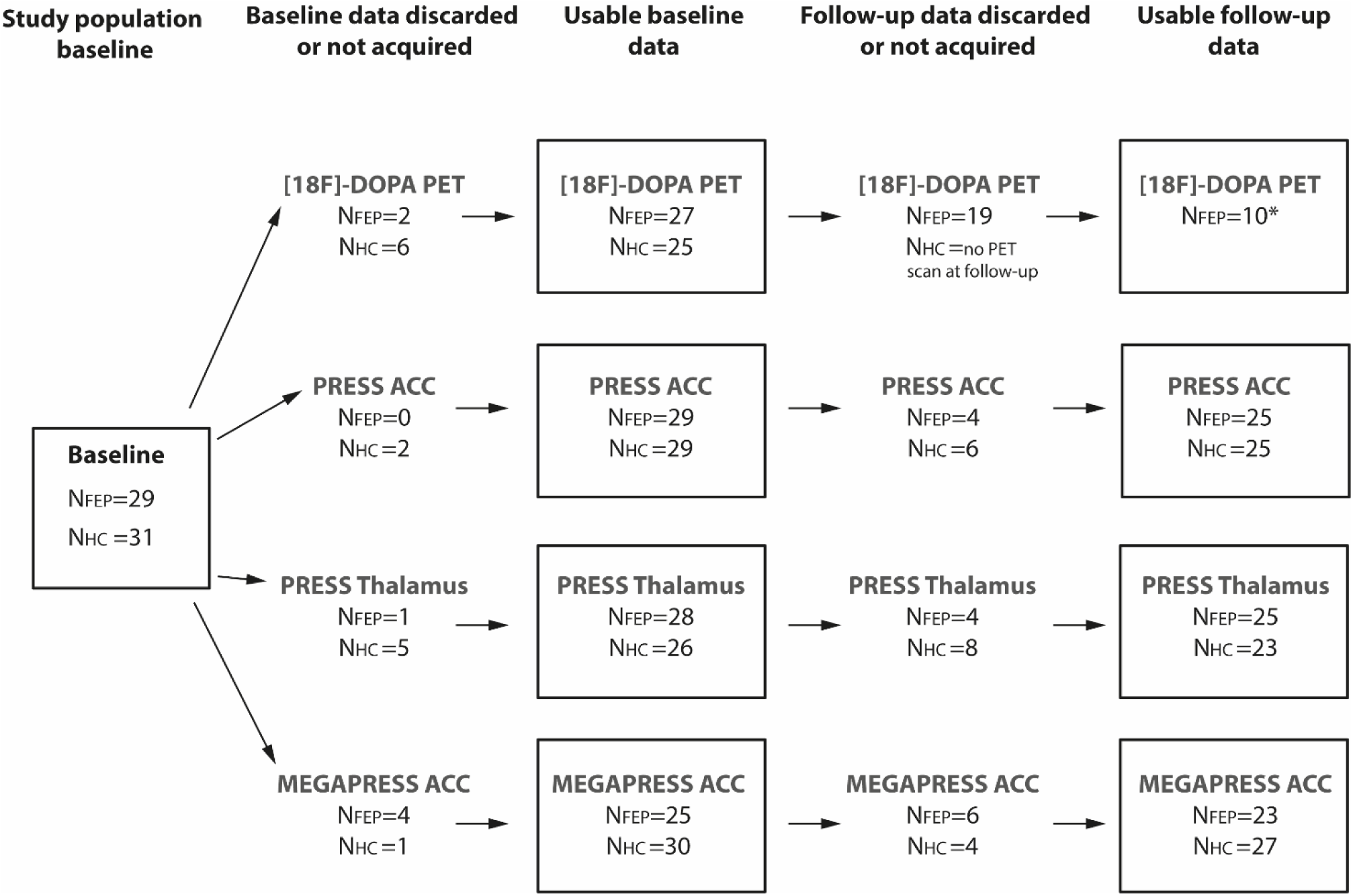
Study flow-chart showing the usable PET and MRS data at baseline and six weeks follow-up visit as well as data that was discarded or not acquired. Abbreviations: [18F]-DOPA PET: 3,4-dihydroxy-6-[^18^F]fluoro-L-phenylalanine; PRESS: point-resolved spectroscopy; MEGAPRESS: Mescher-Garwood point-resolved spectroscopy; ACC: Anterior cingulate cortex; FEP: first-episode patients with psychosis; HC: healthy controls.

Table 1 displays demographic and clinical characteristics. As expected, patients were less educated than HCs, and patients were more frequently smokers. Mean doses and plasma concentrations of aripiprazole are listed. Patients were moderately ill, with baseline PANSS total scores on 78. Patients improved on all PANSS items after treatment as shown in Table 1.

**Table 1:** Clinical and demographic characteristics.

|  | AN-FEP | HC | Statistics |
| --- | --- | --- | --- |
| **Number of participants:** N (F/M) | 29 (19/10) | 31 (20/11) | $\chi^2=0.007$, $p=0.94$ |
| **Number of participants, follow-up, N** | 27 | 30 |  |
| **Age** ± SD, years | 21.9 ± 3.5 | 22.2 ± 3.5 | T(58)=-0.33, $P=0.74$ |
| **Parental socioeconomic status**<br>(high/moderate/low) | 7/18/4 | 10/19/2 | Fisher's $P=0.63$ |
| **Education** ± SD, years | 12.5 ± 2.2 | 14.3 ± 2.6 | T(58)=-2.89, $P=0.005$ |
| **Ethnicity** (n) White/Asian/Middle east | 26/2/1 | 31/0/0 |  |
| **Duration of untreated psychosis**, Median<br>(25–75<sup>th</sup> percentile) in weeks | 25.0<br>(12.0 - 52.0) |  |  |
| **Current tobacco use**, yes/no | 13/16 | 5/26 | $\chi^2=5.88$, $P=0.02$ |
| **Current cannabis use**, yes/no | 2/27 | 1/30 | Fisher's $P=0.61$ |
| **Symptoms at baseline** |  |  |  |
| **PANSS Positive**, mean ± SD | 19.6 ± 3.4 | NA | |
| **PANSS Negative**, mean ± SD | 20.8 ± 5.1 | NA | |
| **PANSS General**, mean ± SD | 37.5 ± 6.2 | NA | |
| **PANSS Total**, mean ± SD | 78.2 ± 11.5 | NA | |
| **Symptoms after six weeks** |  |  |  |
| **PANSS Positive**, mean ± SD | 13.9 ± 3.6 | NA | T(26)=86.5, $P<0.001^*$ |
| **PANSS Negative**, mean ± SD | 16.4 ± 5.2 | NA | T(26)=33.0, $P<0.001^*$ |
| **PANSS General**, mean ± SD | 29.2 ± 7.4 | NA | T(26)=38.6, $P<0.001^*$ |
| **PANSS Total**, mean ± SD | 59.6 ± 12.8 | NA | T(26)=86.0, $P<0.001^*$ |
| **Aripiprazole mean dose at six weeks** ± SD | 9.4 ± 3.7 | NA | |
| **Serum-Aripiprazole level at six weeks** in<br>$\mu\text{g/L}$ ± SD | 124.2 ± 66.8 | NA | |

Abbreviations: AN-FEP: Antipsychotic-naïve patients with first-episode psychosis; HC, healthy controls; N: number; SD, standard deviation; F, female; M, male; PANSS, Positive and Negative Syndrome Scale. \*: Difference between baseline and follow-up score.

### Differences in neurotransmitter interrelations between patients and HC at baseline

The results of the multiple linear regression analyses testing the interrelations between DS in NAcc and GABA levels in ACC, as well as Glu levels in ACC or thalamus are provided in Table 2 and described below.

**Table 2:** Interrelations between dopamine synthesis and levels of GABA and glutamate.

Multiple linear regression models of the relation between dopamine synthesis in Nucleus Accumbens (k<sub>3</sub>) and different combinations of GABA levels in anterior cingulate cortex (ACC), glutamate levels in ACC, and glutamate levels in thalamus in antipsychotic-naïve first-episode patients with psychosis (AN-FEP) and healthy controls (HC).
|  | Group<br>(AN-FEP vs HC) |  | GABA levels in<br>ACC |  | Group*Neuro-<br>transmitter<sup>a</sup> |  | Glutamate levels<br>in thalamus |  | Glutamate levels<br>in ACC |  | Adjusted R-squared (%) |
| --- | --- | --- | --- | --- | --- | --- | --- | --- | --- | --- | --- |
|  | Coef. | p-value | Coef. | p-value | Coef. | p-value | Coef. | p-value | Coef. | p-value |  |
| **Model 1\***<br>k<sub>3</sub> = Group + GABA + Group*GABA + sex | -0.0052 | 0.78 | 0.008 | 0.67 | 0.06 | **0.002** | - | - | - | - | 14.1% |
| **Model 2\***<br>k<sub>3</sub> = Group + GABA + Group*GABA + Glu thal + sex | -0.0011 | 0.96 | 0.01 | 0.64 | 0.06 | **0.007** | 0.0323 | 0.11 | - | - | 17.4% |
| **Model 3**<br>k<sub>3</sub> = Group + Group*GABA + Glu ACC + sex | -0.0069 | 0.73 | 0.01 | 0.63 | 0.06 | **0.003** | - | - | -0.0162 | 0.39 | 13.3% |
| **Model 4**<br>k<sub>3</sub> = Group + Group*Glu thal + Glu thal + sex | -0.0045 | 0.82 | - | - | -0.003 | 0.90 | 0.0417 | **0.04** | - | - | 0.8% |
| **Model 5**<br>k<sub>3</sub> = Group + Group*Glu ACC + Glu ACC + sex | -0.0081 | 0.68 | - | - | 0.03 | 0.16 | - | - | -0.0124 | 0.53 | 2.1% |

<sup>a</sup>Group\*GABA for model 1, 2 and 4; group\*glu thal for model 3; and group\*glu ACC for model 5. \*: p<0.05.
Abbreviations: AN-FEP: antipsychotic-naïve first-episode patients with psychosis; HC: healthy controls; ACC: anterior cingulate cortex; Glu: Glutamate; thal: Thalamus.

#### The interrelation between DS in NAcc and GABA levels in ACC

DS was modelled by treating k_3_ in NAcc as the dependent variable and group, GABA levels, and the group-GABA interaction as independent variables (Table 2, Model 1). This model was significant and driven by the group-GABA interaction (p=0.002) indicating an inverse relationship between GABA levels and k_3_ in AN-FEP compared to HC, as hypothesized. Post hoc analyses revealed a positive association between GABA in ACC and dopamine activity in NAcc in patients (p=0.04, β=0.15) and a negative association in HC (p=0.03, β=-0.15) (Figure 2A).

**Figure 2:**
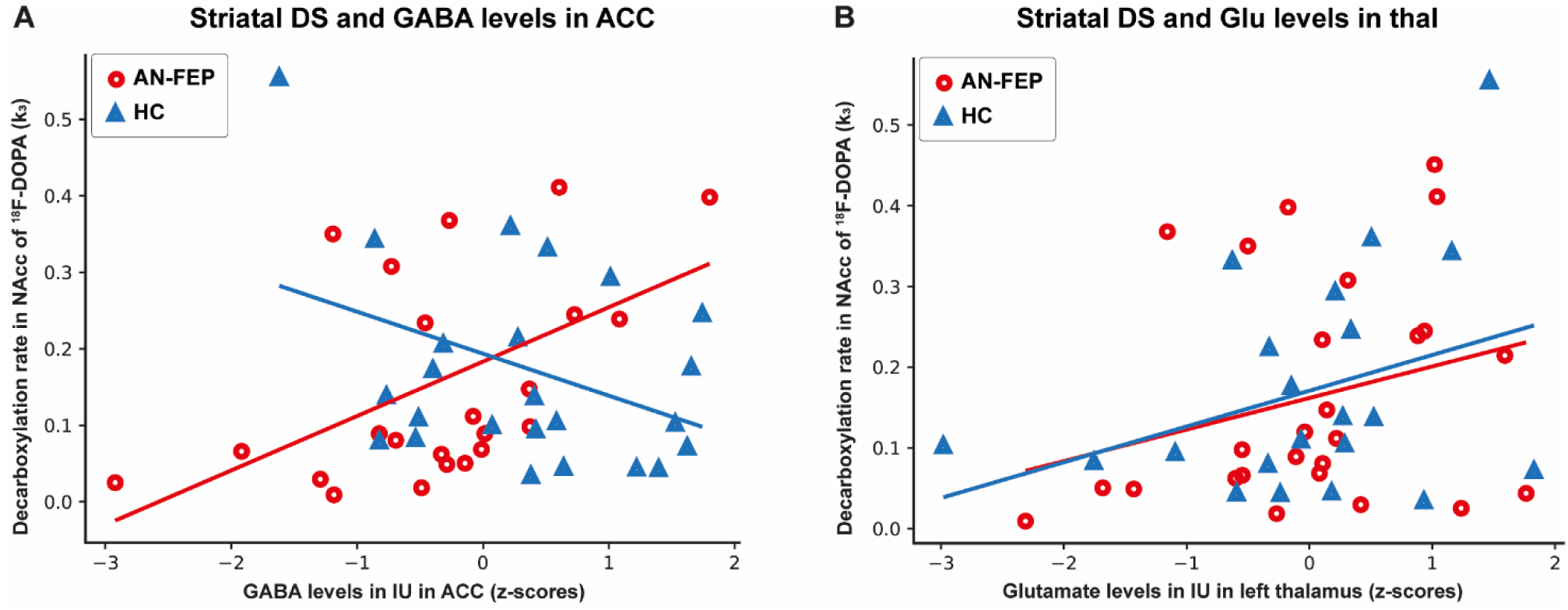
The interrelation between decarboxylation rate k_3_ in nucleus accumbens (NAcc) and GABA levels in anterior cingulate cortex (ACC) (Table 2, Model 1) (A) as well as glutamate levels in left thalamus (Table 2, Model 3) (B) in antipsychotic-naïve patients with first-episode psychosis (AN-FEP) (red circles) and healthy controls (HC) (blue triangles). A: An inverse relationship between k_3_ in NAcc and GABA levels in ACC was observed in AN-FEP compared with HC (p=0.002) due to a positive association in patients (p=0.04, β =0.15) but a negative in HC (p=0.03, β =-0.15). B: A positive association was found between k_3_ in NAcc and glutamate levels in thalamus in both AN-FEP and HC (p=0.04, b=0.04). Abbreviations: DS: dopamine synthesis; NAcc: nucleus accumbens; ACC: anterior cingulate cortex; AN-FEP: antipsychotic-naïve patients with first-episode psychosis; HC: Healthy controls; IU: Institutional units.

When examining combinations of the independent variables GABA in ACC, Glu in ACC, and Glu in thalamus we found that a combination of GABA in ACC and Glu in thalamus was significantly related to k_3_ (Table 2, Model 2). The association was driven by the group*GABA interaction (p=0.007) and not GABA in ACC or Glu in thalamus. Similarly, a combination of GABA in ACC and Glu in ACC revealed a significant association between k_3_ and the group*GABA interaction (p=0.003) whereas GABA in ACC and Glu in ACC were insignificant (Table 2, model 3).

#### The interrelation between DS in NAcc and Glu in either thalamus or ACC

Explorative analyses of the association between k_3_ and levels of either Glu in thalamus or Glu in ACC revealed a significant positive association between k_3_ and thalamic glutamate levels that did not differ between AN-FEP and HC (Table 2, model 4 and Figure 2B), whereas there were no significant associations between k_3_ and ACC glutamate levels (Table 2, Model 5, Supplementary Figure S2). Sex was included as a covariate but did not contribute to any of the models.

### The relation between neurotransmitter interrelations and improvement in positive symptoms

Multiple linear regression models examining the relation between improvement in positive symptoms after six weeks of treatment with aripiprazole and striatal DS as well as glutamate and GABA levels are described below and summarized in Supplementary Table S5.

First, we tested the second hypothesis that an abnormal relationship between striatal DS and GABA levels in ACC had an impact on treatment response. Improvement in positive psychotic symptoms was associated with k_3_ (p=0.021) and the GABA-k_3_ interaction (p=0.023). The GABA-k_3_ interaction indicated that having either high or low values of both k_3_ and GABA at baseline were associated with a better treatment response (i.e., negative value of ΔPANSS positive), whereas low GABA and high k_3_ values, or the opposite, were associated with a poor treatment response (Figure 3A and Supplementary Table S5, Model 6).

**Figure 3:**
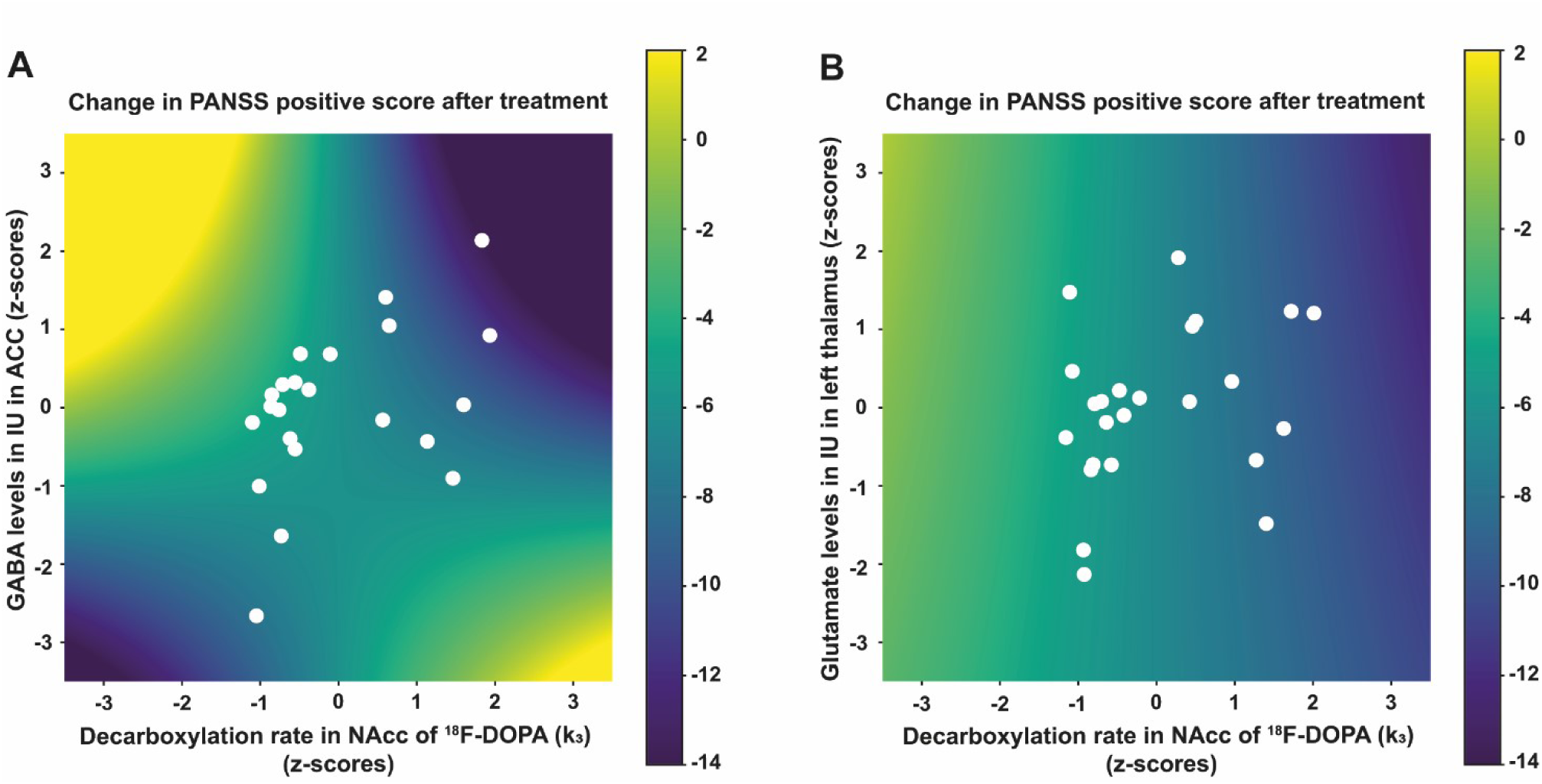
Relations between treatment response, striatal DS, and metabolite levels. The effect of k_3_ (NAcc) and either GABA levels in ACC (A) or Glu levels in thalamus (B) and their interaction on the treatment response (change in PANSS positive). The colorbars indicate the change in PANSS positive score after treatment, where a positive change indicates worsening symptoms, and a negative change indicates improvement. The white dots illustrate the individual patients. The neurotransmitters are standardized to zero mean and unit variance. A) Illustrates the significant interaction between k_3_ and GABA in ACC. If both k_3_ and GABA are either low or high, the patient improves during the six weeks of treatment (dark blue). In contrast, if GABA is high and k_3_ is low or opposite, the patient has more symptoms after treatment (yellow). B) Illustrates a significant effect of k_3_ on the treatment response, but no interaction effect between k_3_ and Glu in thalamus: the treatment response only depends on the value of k_3_, i.e., the color only changes along the x-axis and not along the y-axis.

When GABA levels were exchanged with Glu levels in thalamus (Supplementary Table S5, Model 7), the overall model was significant, but this was driven by a significant relation between treatment response and k_3_ (p=0.008) as well as sex (p=0.015) (due to better treatment response in male patients), whereas the relation between treatment response and the k_3_*Glu in thalamus interaction was insignificant, suggesting that Glu in thalamus was not related to treatment response. Similar findings were observed when GABA levels were exchanged with Glu levels in ACC (Supplementary Table S5, Model 8), where the overall model was significant driven by a significant relation between treatment response and k_3_ (p=0.015) but not Glu in ACC or the k_3_*Glu in ACC interaction.

### Changes in neurotransmitter interrelations after treatment

At baseline, there was a significant positive association between k_3_ and Glu in thalamus in the patients (p=0.002) (as also previously illustrated in figure 2B) that decreased over time (p=0.013) (Figure 4). We did not observe significant changes after treatment in the interrelation between k_3_ and GABA (p=0.31) or in the interrelation between k_3_ and Glu levels in ACC (p=0.73) in the patients (Supplementary Figures S3 and S4).

**Figure 4:**
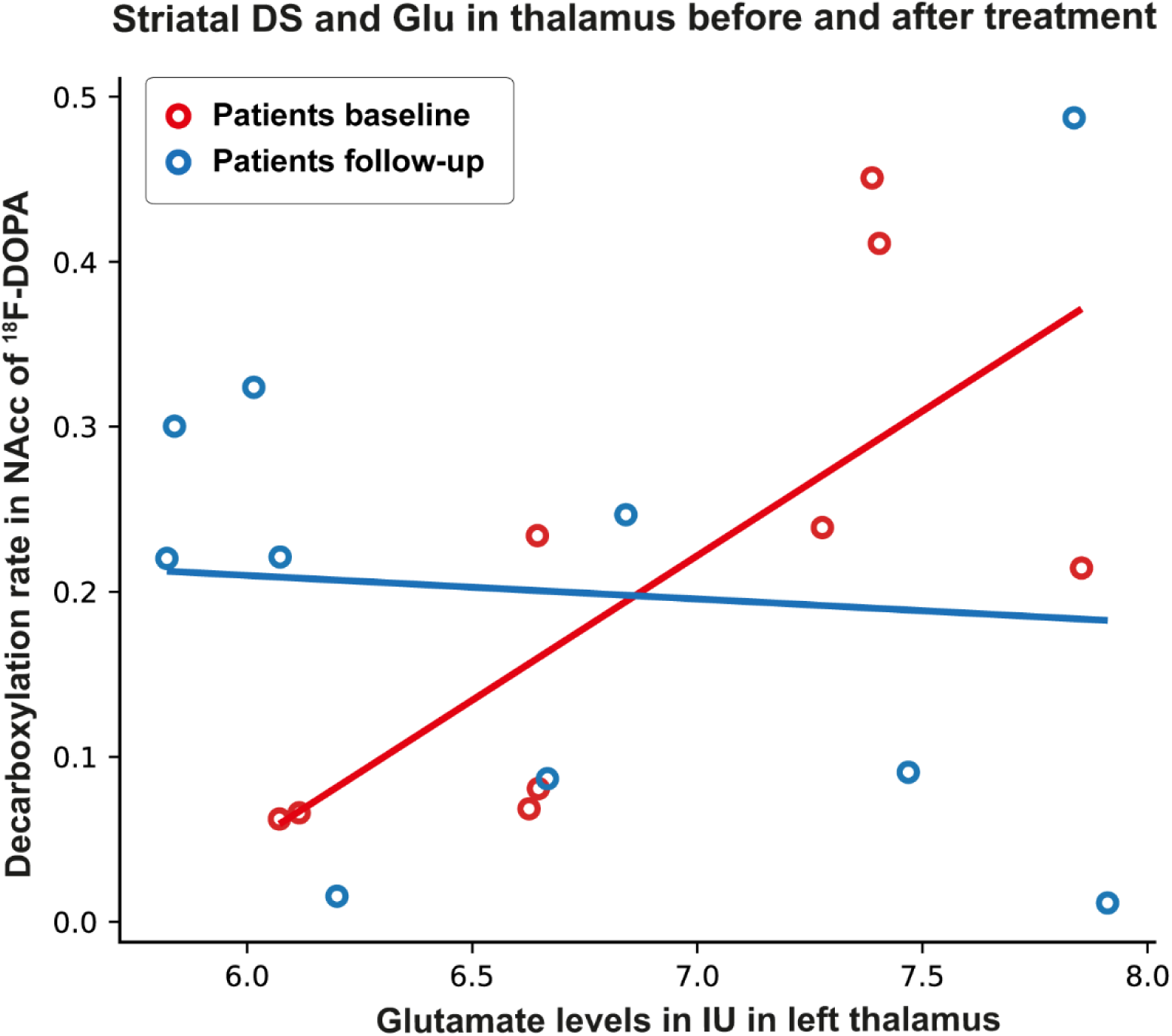
Figure 4 illustrates that the association between k_3_ in NAcc and glutamate levels in thalamus decreased from before (red circles) to after 6 weeks of treatment with aripiprazole (blue circles) in the subgroup of initially antipsychotic-naïve patients with first-episode psychosis assessed both before and after treatment (Glu in thalamus*visit: p=0.013). Abbreviations: DS: dopamine synthesis; NAcc: nucleus accumbens; ^18^F-DOPA: 3,4-dihydroxy-6-[^18^F]fluoro-L-phenylalanine; IU: Institutional units.

## Discussion

To our knowledge, the present study is the first to characterize the interrelations between DS in NAcc, cortical GABA levels, and cortical and thalamic Glu levels in AN-FEP compared with matched HC and to relate these interrelations to treatment response in the patients. The data confirmed our main hypotheses, i.e., that GABA levels in ACC and DS in NAcc are inversely interrelated in AN-FEP and HC, and that the degree of abnormality is associated with treatment response. In addition, we found a significant positive relationship between Glu levels in thalamus and k_3_ in both patients and HC. The relationship between Glu levels in thalamus and k_3_ changed in the patients after treatment but was not associated with treatment response. We found no significant group differences in the interrelations between k_3_ and Glu levels in ACC nor any changes in the relationship in the patients after treatment.

The finding of an inverse interrelation between striatal dopaminergic and cortical GABAergic activity in AN-FEP and HC support previous data showing that this interrelation is pivotal in the separation of patients from HC (24), and further extend these findings by revealing that the abnormality in patients is a positive instead of negative association between ACC GABA levels and striatal dopaminergic activity. Interestingly, dysfunction of cortical GABAergic interneurons has long been hypothesized to be implicated in the development of psychosis (39, 40). Development of prefrontal cortex in adolescence is assumed to be initiated by maturation of inhibitory GABAergic function (41–46) and the meso-cortical dopaminergic system (47, 48). For schizophrenia, it is suggested that abnormalities in this neurodevelopmental period may contribute to development of psychopathology (47), which is in line with the present findings showing an abnormal relationship between striatal dopaminergic activity and prefrontal GABAergic levels at illness onset in AN-FEP.

The mechanisms underlying the dysfunctional interrelations between cortical GABAergic and subcortical dopaminergic activity remains to be clarified, but preclinical studies have shown that GABAergic interneurons control the output of glutamatergic pyramidal cells believed to have a downstream effect on among others striatal dopaminergic activity via the direct cortico-striato-thalami-cortical pathway – or the accelerator pathway – and the indirect so-called brake pathway (2, 6, 49). According to this model, both increased and decreased cortical glutamatergic activity can increase subcortical dopaminergic activity via the direct, respectively the indirect pathway, and normal information-processing depends on a balance between the two pathways. In line with this, the present data show a negative relationship between GABA levels in ACC and DS in NAcc in HC, supporting the notion that striatal DS is dampened by increased cortical GABA activity in normal conditions, whereas the relationship is deviant in the AN-FEP.

The inverse relationship between ACC GABA levels and striatal DS is not, however, in line with our earlier findings of a negative association between cortical GABAergic levels and striatal perfusion in HC as well as AN-FEP (16). However, perfusion reflects activity of other neurotransmitters than dopaminergic (50) and more precise measures of striatal activity are probably needed to reveal abnormalities in the prefrontal GABA-striatal dopamine interrelation at illness onset.

Next, we explored if combined measures of GABA levels in ACC and Glu in thalamus and ACC could characterize the variation in striatal DS in AN-FEP more accurately than GABA levels alone. We found that a combination of GABA in ACC, Glu in thalamus and group*GABA improved accuracy of explaining variation in striatal DS (Table 2, model 2) although it was only the group*GABA interaction that was significantly associated with k_3_. This is in line with previous findings in an overlapping sample showing that AN-FEP could be separated from HC with higher precision when combining measures of striatal DS, GABA in ACC, Glu in thalamus and the GABA*striatal DS interaction, but that only the GABA*striatal DS interaction was a significant predictor of patients status (24). The findings commonly suggest that the abnormal GABA*striatal DS interaction is central to the schizophrenia pathophysiology.

Although Glu in thalamus did not significantly explain the variance in striatal DS, we found a significant positive association between DS in striatum and thalamic Glu levels that did not differ between AN-FEP and HC (Figure 2B). This corresponds to a previous study showing a positive association between subcortical DS and Glu levels in HC (23). The data suggest that subcortical DS and Glu are related, but not disturbed in AN-FEP.

Notably, we did not find a significant negative association between presynaptic striatal dopamine activity and Glu levels in ACC in the antipsychotic-naïve patients with first-episode psychosis (Supplementary Figure S2), nor changes in this relationship after treatment (Supplementary Figure S3 and Table S5, Model 8). This seemingly contrasts earlier findings by Jauhar and colleagues (11, 28), but the present data do in many ways differ from the data included in the two previous publications. First, all patients in the present study were strictly antipsychotic-naïve and within the schizophrenia spectrum and both medication status and diagnosis might influence neurotransmitter activity. Second, in the present study the patients received six weeks of antipsychotic monotherapy with the partial D_2_ receptor agonist aripiprazole compared with naturalistic treatment (28). Third, we placed our ACC voxel more rostral where Glu levels are lower compared to ventral ACC in preclinical and HC studies (26, 51–53). Fourth, we used arterial blood sampling and a 4P PET model allowing us to measure the striatal decarboxylation rate of [18F]-DOPA to [18F]-dopamine (k_3_) in contrast to Jauhar et al. who used the traditional TR method to assess DSC (K_i_). We chose k_3_ as our dependent variable since we have previously found it to be superior to K_i_ assessed with both the TR and the 4P method (7). Last, only 21% of the patients in Jauhar et al. 2018 (11) were female in contrast to 66 % in the present study and ethnic differences were present as well.

In line with our second hypothesis, both the GABA-DS interrelation and DS were predictors for treatment response (Figure 3 and Supplementary Table S5, Model 6). The data suggest that the effect on psychotic symptoms is related to either low or high values of both GABA and k_3_, i.e., the more the interrelation in the patients differ from that seen in HC, the better the response to treatment with the partial D_2_ receptor agonist, aripiprazole. We did not, however, see a significant change in the association between k_3_ and GABA after treatment, but Supplementary Fig. S4 could suggest that this might be due to loss of patients to follow-up. Preclinical studies indicate several regulatory mechanisms between the dopaminergic and GABAergic neurotransmitter systems. For example, dopamine D_1_ and D_2_ as well as D_4_ receptors are present on cortical GABAergic interneurons, and timing, strengths of synaptic input, and firing state of the dopaminergic neurons likewise modulate GABAergic interneurons (3, 4, 54–56). Future research should aim at unravelling these mechanisms further, as this may pave the way for development of novel treatment targeting a disturbed GABA-DS interrelation (57).

We did not find that combined measures of Glu in thalamus and DS or Glu in ACC and DS were associated with treatment response after six weeks (Supplementary Table S5, Model 7 and 8) although previous studies of single neurotransmitters (studied separately) in overlapping cohorts have shown an association between Glu in thalamus in AN-FEP and short-term response as well as between decreased GABA levels in ACC at illness onset and outcome after both short and longer term treatment (16, 19). These discrepancies in findings may reflect a power issue as the current study-sample was smaller than in the previous studies since DS in striatum is technically more challenging to measure.

Last, we explored if treatment changed the interrelations between striatal DS, GABA and glutamate levels. We found that the interrelation between k_3_ and thalamic Glu changed after six weeks of treatment with aripiprazole, but the change was not related to treatment response. A change was expected given that we have earlier found that Glu levels in thalamus changes after treatment (19), whereas k_3_ in NAcc did not (7). We did not observe significant changes in neither the interrelation between k_3_ and GABA nor between k_3_ and Glu in ACC after treatment, which is in line with previous studies of individual neurotransmitter where we neither observed changes in k_3_, nor in GABA levels in ACC, or Glu in ACC after six weeks treatment (7, 19). As afore mentioned, loss to follow-up of k_3_ measures might also explain why no significant changes in the interrelation between k_3_ in NAcc and GABA levels in ACC were found despite a visual interpretation of a change after treatment (Supplementary Figure S4), whereas this does not seem to be the case for the negative findings regarding changes in the interrelation between k_3_ in NAcc and Glu levels in ACC (Supplementary Figure S3). The findings may suggest that short-term treatment primarily affect glutamate in subcortical regions, whereas cortical regions may be more affected after longer-term treatment (16).

Three major strengths of the current study are inclusion of strictly antipsychotic-naïve patients, measures of striatal DS using an arterial input function thereby allowing us to measure the decarboxylation rate of [18F]-DOPA to [18F]-dopamine (k_3_), and the combined measures of both striatal DS, GABA as well as glutamate levels. Moreover, AN-FEP were free from substance abuse and acute effects of benzodiazepines, carefully matched to HC, and studied in a longitudinal design. However, limitations should also be addressed. First, MRS measures of Glu and GABA cannot differentiate between extracellular and intracellular GABA and Glu concentrations. Moreover, the sample size for k_3_ at follow-up is limited given the technical challenges with striatal DS measures.

### Conclusion and future directions

The present study characterizes as the first aberrant interrelations between DS in NAcc and GABA levels in ACC in AN-FEP that predict the effect of subsequent antipsychotic treatment. We additionally found identical significant interrelations between DS in NAcc and Glu levels in thalamus in patients and HC. The interrelation between k_3_ and thalamic Glu levels changed over time in the patients but was not associated with treatment response. The pathophysiological mechanisms behind the observed, clinically relevant, abnormalities in the interrelations between cortical GABAergic and subcortical dopaminergic activity, as well as their implications for future treatment strategies, should be further studied in future longitudinal studies on AN-FEP involving additional modalities such as functional and structural connectivity and brain structure as well as additional clinical data on among others cognitive functions and early information processing.

## Funding and acknowledgements

This study was funded by an independent grant from the Lundbeck Foundation (R155-2013-16337) to the Lundbeck Foundation Centre of Excellence for Clinical Intervention and Neuropsychiatric Schizophrenia Research (CINS) (Glenthøj), grants from the Wørzner and Gerhard Linds Foundations and support from the Mental Health Services, Capital Region of Denmark (Glenthøj and Ebdrup). PhD grants and a post doc grant were obtained from the Mental Health Services in the Capital Region of Denmark (Sigvard), the Faculty of Health and Medical Sciences, University of Copenhagen (Bojesen), the research Fund 2022 in the Capital Region of Denmark (Bojesen), and an independent grant from the Lundbeck Foundation (R403-2022-1361). The funding sources had no role in the design or conduction of the study design, nor in the collection, analyses, and interpretation of data, or in the writing, review approval and submission of the manuscript for publication. We further wish to thank the staff at Center for Neuropsychiatric Schizophrenia Research (CNSR) and the Centre for Clinical Intervention and Neuropsychiatric Schizophrenia Research (CINS), Mental Health Centre Glostrup, University of Copenhagen; the PET Centre, Herlev Hospital, University of Copenhagen and Functional Imaging Unit, Rigshospitalet, Glostrup for help.

## Clinical trial registration

The Pan European Collaboration on Antipsychotic Naïve Schizophrenia II (PECANSII) study, ClinicalTrials.gov Identifier: NCT02339844. https://www.clinicaltrials.gov/study/NCT02339844

## Conflict of Interest

Dr. Glenthøj has been the leader of a Lundbeck Foundation Centre of Excellence for Clinical Intervention and Neuropsychiatric Schizophrenia Research (CINS) (January 2009 – December 2021), which was partially financed by an independent grant from the Lundbeck Foundation based on international review and partially financed by the Mental Health Services in the Capital Region of Denmark, the University of Copenhagen, and other foundations. All grants are the property of the Mental Health Services in the Capital Region of Denmark and administrated by them. She has no other conflicts to disclose.

Dr. Bojesen received lecture fees from Lundbeck Pharma A/S. Dr. Ebdrup BE is part of the Advisory Board of Boehringer Ingelheim, Lundbeck Pharma A/S; and has received lecture fees from Boehringer Ingelheim, Otsuka Pharma Scandinavia AB, and Lundbeck Pharma A/S.

The rest of the authors have no conflicts of interest to disclose.

## Data Availability

First authors can be contacted about availability of the data.

## Notes

### Competing Interest Statement

Dr. Glenthoej has been the leader of a Lundbeck Foundation Centre of Excellence for Clinical Intervention and Neuropsychiatric Schizophrenia Research (CINS) (January 2009 to December 2021), which was partially financed by an independent grant from the Lundbeck Foundation based on international review and partially financed by the Mental Health Services in the Capital Region of Denmark, the University of Copenhagen, and other foundations. All grants are the property of the Mental Health Services in the Capital Region of Denmark and administrated by them. She has no other conflicts to disclose. Dr. Bojesen received lecture fees from Lundbeck Pharma A/S. Dr. Ebdrup BE is part of the Advisory Board of Boehringer Ingelheim, Lundbeck Pharma A/S; and has received lecture fees from Boehringer Ingelheim, Otsuka Pharma Scandinavia AB, and Lundbeck Pharma A/S.
The rest of the authors have no conflicts of interest to disclose.

### Clinical Protocols

https://www.clinicaltrials.gov/study/NCT02339844

### Author Declarations

The Committee on Biomedical Research Ethics in the capital region of Denmark gave ethical approval for this work (H-3-2013-149)

