## Supplementary information for "Interrelations Between Dopaminergic-, GABAergic- and Glutamatergic Neurotransmitters in Antipsychotic-Naïve Psychosis Patients and the Association to Initial Treatment Response"

### Supplementary Methods

*Minimum reporting standards for in vivo magnetic resonance spectroscopy*

Minimum reporting standards for the magnetic resonance spectroscopy are provided in the supplementary Table S1 (1).

**Supplementary Table S1: Minimum reporting standards for Magnetic Resonance Spectroscopy**

| | |
|---|---|
| Site (name) | Center for Neuropsychiatric Schizophrenia Research (CNSR), Mental Health Center Glostrup, DK |
| 1. Hardware | |
| a. Field strength [T] | 3T |
| b. Manufacturer | Philips Achieva |
| c. Model (software version if available) | Philips dSTREAM Achieva |
| d. RF coils: nuclei (transmit/receive), number of channels, type, body part | 32-channel head coil |
| e. Additional hardware | None |
| 2. Acquisition | |
| a. Pulse sequence | PRESS and MEGAPRESS |
| b. Volume of interest (VOI) locations | Voxel locations are shown in Figure S1:<br>Dorsal Anterior cingulate cortex (dACC) for MEGAPRESS acquisitions (Supplementary Figure S1A)<br>Left thalamus and dACC for PRESS acquisitions (Supplementary Figure S1B and C) |
| c. Nominal VOI size [cm3] | ACC: 3.0*3.0*3.0cm$^3$<br>Left thalamus: 2.0*1.5*2.0cm$^3$ |
| d. Repetition time (TR), echo time (TE) [ms] | MEGAPRESS: TR=2000ms, TE=68ms<br>PRESS: TR=3000ms, TE=30ms |

| | |
|---|---|
| e. Total number of excitations or acquisitions per spectrum | MEGAPRESS: 320 averages<br><br>PRESS: 128 averages |
| f. Additional sequence parameters (spectral width in Hz, number of spectral points, frequency offsets) | 2500 Hz, 1024 samples |
| g. Water suppression method | MOIST |
| h. Shimming method, reference peak, and thresholds for “acceptance of shim” chosen | Second order Pencil Beam auto |
| i. Triggering or motion correction method (respiratory, peripheral, cardiac triggering, incl. device used and delays) | None |
| 3. Data analysis methods and outputs | |
| a. Analysis software | LCModel version 6.3-1L for PRESS acquisitions<br><br>Gannet version 3.1 for MEGAPRESS acquisitions |
| b. Processing steps deviating from quoted reference or product | None |
| c. Output measure (eg absolute concentration, institutional units, ratio), processing steps deviating from quoted reference or product | Metabolite concentrations adjusted for partial-volume cerebrospinal fluid and the fraction of gray and white matter in the voxel.<br><br>No processing steps deviated from quoted references or product. |
| d. Quantification references and assumptions, fitting model assumptions | LCModel basis set: press_te30_3t_gsh_v3<br><br>Gannet uses nonlinear regression for signal model fitting, with fit parameters optimized using the least-squares Levenberg-Marquardt algorithm. For increased computational speed and a better solution, the starting values of the optimization are derived from a “pre-fit” that uses the trust-region-reflective algorithm. Metabolite values are quantified using either the 3.0 ppm total creatine or unsuppressed water signal (water signal was used as quantification reference in the present study). |

| 4. Data quality | |
|---|---|
| a. Reported variables (SNR, linewidth (with reference peaks)) | SNR and FWHM are reported for PRESS acquisitions in Supplementary Table S2 and FWHM for MEGAPRESS acquisitions in Supplementary Table S3 |
| b. Data exclusion criteria | First, visual inspection was used to exclude spectra.<br>For PRESS acquisitions, individual metabolite values with a Cramér-Rao lower bound (CRLB) > 20% as reported by LCModel were excluded.<br>For MEGAPRESS data, individual GABA values with signal fit error >15% as reported by Gannet was excluded |
| c. Quality measures of postprocessing model fitting (eg CRLB, goodness of fit, SD of residual) | CRLB (Supplementary Table S2) for PRESS acquisitions<br>Goodness of fit (Supplementary Table S3) for MEGAPRESS acquisitions |
| d. Sample spectrum | Representative spectra are shown in Figure S1 for MEGAPRESS acquisitions in dACC (D), PRESS acquisitions in left thalamus (E), and PRESS acquisitions in dACC (F). |

**Supplementary Figure S1**

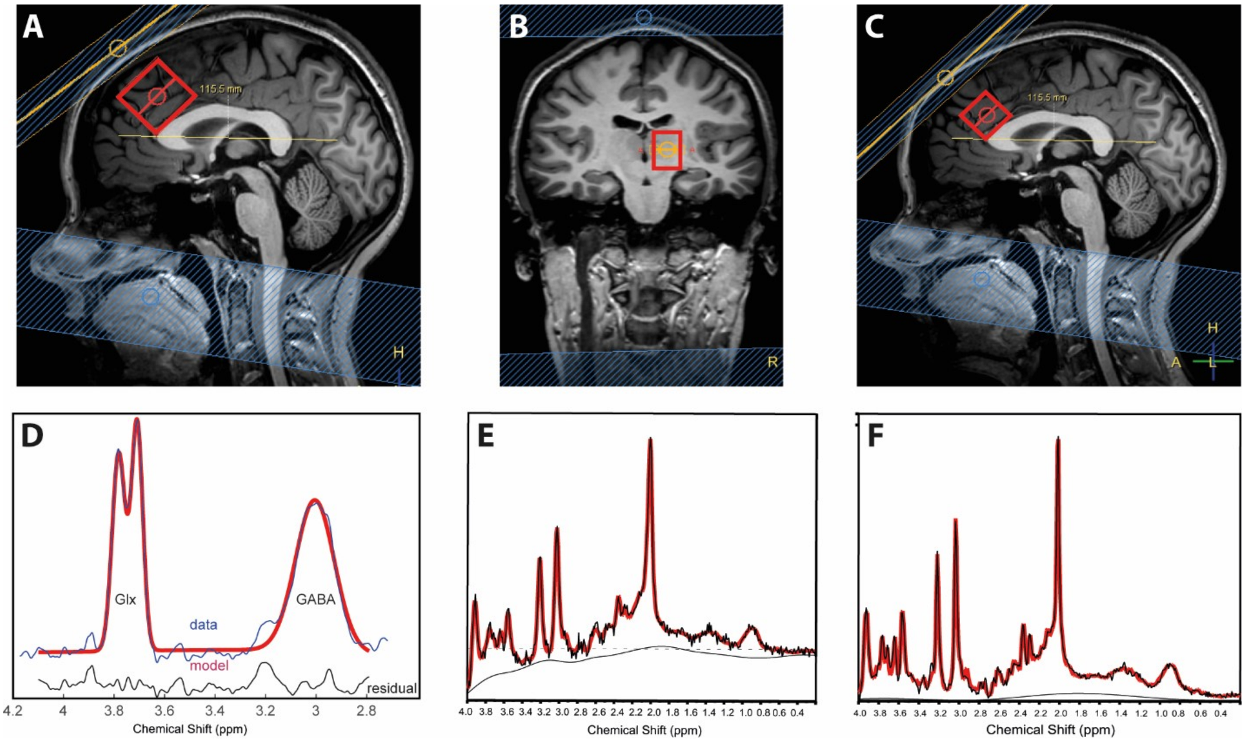


Supplementary Figure S1: Voxel location for MEGAPRESS dorsal anterior cingulate cortex (dACC) acquisitions (A) as well as representative spectrums (D), and voxel locations for PRESS acquisitions in left thalamus (B) and dACC (C) together with representative spectra from left thalamus (E) and dACC (F). Black and blue lines in the spectra represents the obtained data and read lines represent the fit made by Gannet and LCModel.
Abbreviations: ppm: parts per million.

**Supplementary Table S2: Data quality for PRESS acquisitions in left thalamus**

| | FEP | HC | Statistics |
|---|---|---|---|
| **PRESS acquisitions in left thalamus baseline** | | | |
| **N** | n=28 | n=26 | |
| FWHM in ppm | 0.048±0.006 | 0.047±0.007 | P=0.66 |
| SNR | 16.4±3.7 | 17.0±3.2 | P=0.55 |
| CRLB (%) Glu | 10.2±1.9 | 10.0±2.1 | P=0.71 |
| **PRESS acquisitions in left thalamus after six weeks** | | | |
| **N** | n=25 | n=23 | |
| FWHM in ppm | 0.048±0.008 | 0.046±0.004 | P=0.28 |
| SNR | 17.2±3.7 | 16.8±3.9 | P=0.77 |
| CRLB (%) Glu | 10.0±1.6 | 9.8±1.5 | P=0.67 |

Supplementary Table S2 shows data quality parameters for PRESS acquisitions in left thalamus. Abbreviations: FEP: First episode psychosis patients; HC: Healthy controls; FWHM: Full-width half-maximum; ppm: parts per million; SNR: Signal-to-noise ratio; CRLB: Cramér-Rao lower bound; Glu: Glutamate

**Supplementary Table S3: Data quality for PRESS acquisitions in anterior cingulate cortex**

| | **FEP** | **HC** | **Statistics** |
|---|---|---|---|
| **PRESS acquisitions in dorsal anterior cingulate cortex baseline** | | | |
| **N** | n=29 | n=29 | |
| FWHM in ppm | 0.029±0.005 | 0.025±0.006 | P=0.01 |
| SNR | 32.3±2.2 | 33.1±2.1 | P=0.12 |
| CRLB (%) Glu | 5.1±0.3 | 5.0±0.5 | P=0.30 |
| **PRESS acquisitions in dorsal anterior cingulate cortex after six weeks** | | | |
| **N** | n=25 | n=25 | |
| FWHM in ppm | 0.026±0.005 | 0.027±0.005 | P=0.68 |
| SNR | 32.0±2.4 | 32.6±2.3 | P=0.23 |
| CRLB (%) Glu | 5.2±0.6 | 5.0±0.2 | P=0.30 |

Supplementary Table S3 shows data quality parameters for PRESS acquisitions in anterior cingulate cortex. Abbreviations: FEP: First episode psychosis patients; HC: Healthy controls; FWHM: Full-width half-maximum; ppm: parts per million; SNR: Signal-to-noise ratio; CRLB: Cramér-Rao lower bound; Glu: Glutamate

**Supplementary Table S4: Data quality for MEGAPRESS acquisitions**

| | **FEP** | **HC** | **Statistics** |
|---|---|---|---|
| **MEGAPRESS acquisitions in anterior cingulate cortex at baseline** | | | |
| **N** | n=25 | n=30 | |
| FWHM in HZ | 8.8±0.6 | 8.5±0.4 | P=0.04 |
| Fit error (%) GABA | 5.7±1.5 | 5.0±1.1 | P=0.03 |
| **MEGAPRESS acquisitions in anterior cingulate cortex after six weeks** | | | |
| **N** | n=23 | n=27 | |
| FWHM in HZ | 8.7±0.5 | 8.6±0.4 | P=0.34 |
| Fit error (%) GABA | 5.0±1.4 | 4.5±0.9 | P=0.11 |

Supplementary Table S4 shows data quality parameters for MEGAPRESS acquisitions in anterior cingulate cortex. Abbreviations: FEP: First episode psychosis patients; HC: Healthy controls; FWHM: Full-width half-maximum; HZ: Hertz; GABA: gamma-aminobutyric acid.

**Supplementary Figure S2**

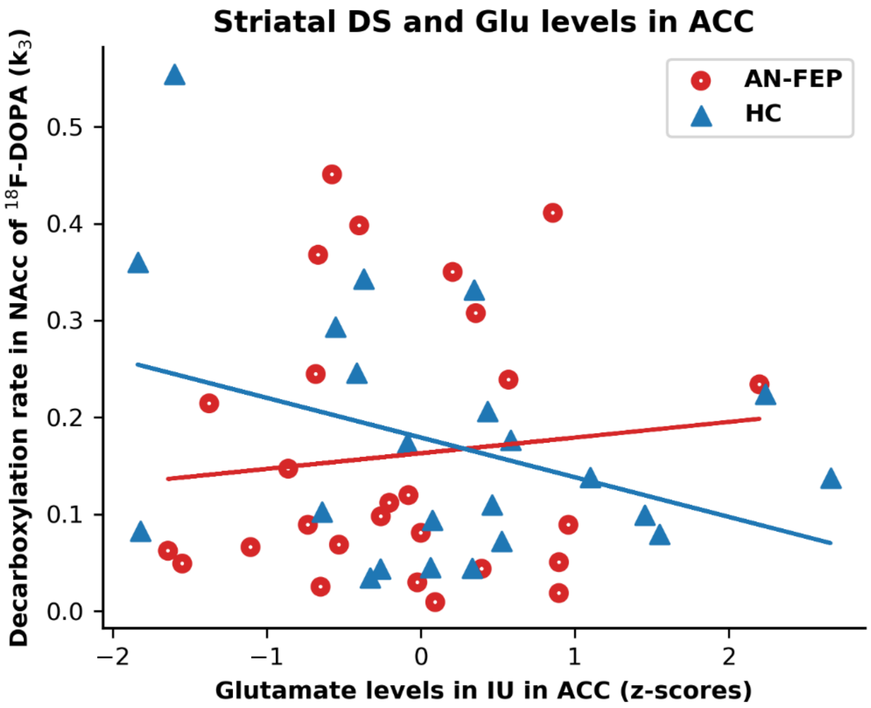


Supplementary Figure S2: The interrelation between decarboxylation rate $k_3$ in nucleus accumbens (NAcc) and Glu levels in anterior cingulate cortex (ACC) in antipsychotic-naïve patients with first-episode psychosis (AN-FEP) (red circles) as well as healthy controls (HC) (blue triangles). Neither the overall model (p=0.56) nor the interaction between group and Glu in ACC (p=0.16) were significant. Abbreviations: NAcc: nucleus accumbens; ACC: anterior cingulate cortex; AN-FEP: antipsychotic-naïve patients with first-episode psychosis; HC: Healthy controls; IU: Institutional units.

**Supplementary Figure S3**

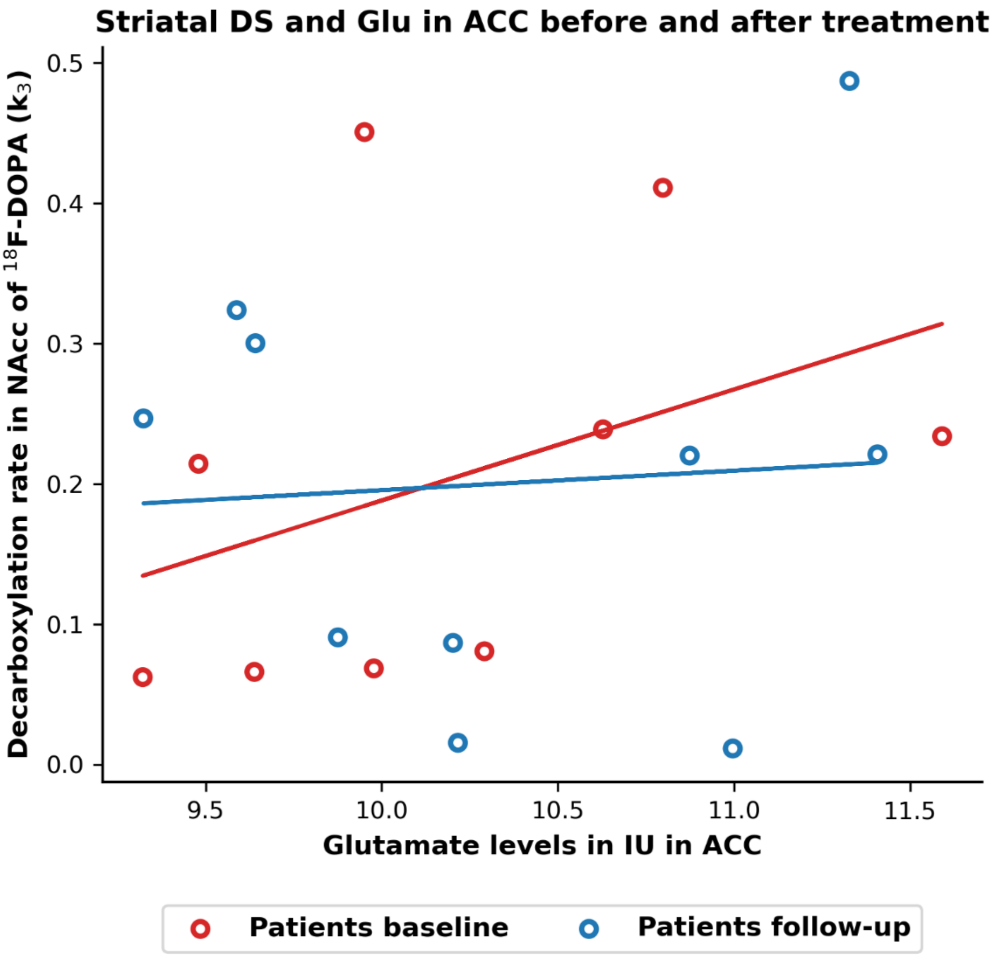


Supplementary Figure S3: The association between $k_3$ in NAcc and glutamate levels in anterior cingulate cortex (ACC) before and after 6 weeks of treatment with aripiprazole in the subgroup of initially antipsychotic-naïve patients with first-episode psychosis assessed both before and after treatment. At baseline, the association between $k_3$ in NAcc and glutamate levels in ACC was insignificant (red circles, $p=0.32$) and this did not change significantly after treatment (blue circles, $p=0.73$). The baseline association in the total group of patients appears from Figure S2. Abbreviations: NAcc: nucleus accumbens; ACC: anterior cingulate cortex; IU: Institutional units.

**Supplementary Figure S4**

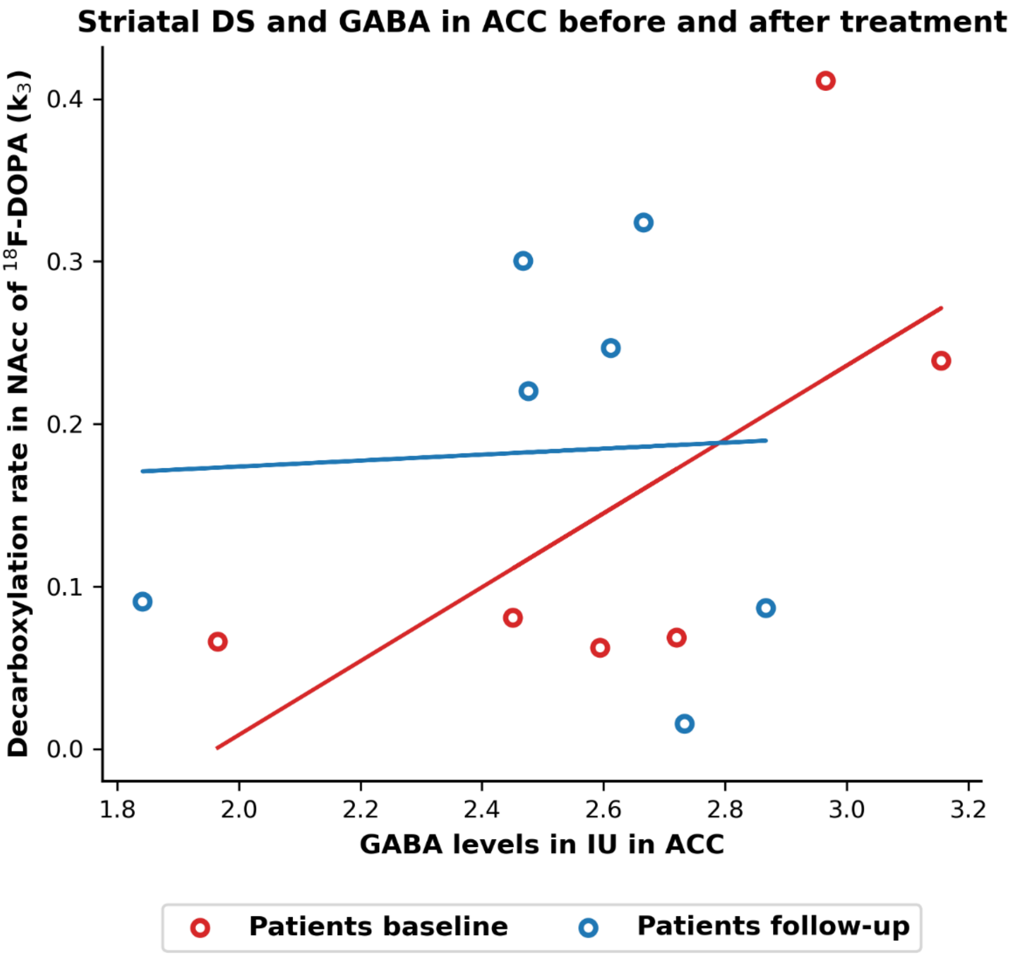


Supplementary Figure S4: The association between $k_3$ in NAcc and GABA levels in anterior cingulate cortex (ACC) before and after 6 weeks of treatment with aripiprazole in the subgroup of initially antipsychotic-naïve patients with first-episode psychosis assessed both before and after treatment. In this subgroup of patients, neither the association between $k_3$ in NAcc and GABA levels in ACC at baseline (red circles, β=0.66, p=0.14) nor the change after treatment (blue circles, β=-0.64, p=0.31) were significant. The baseline association in the total group of patients was significant (Figure 2A). Abbreviations: NAcc: nucleus accumbens; ACC: anterior cingulate cortex; IU: Institutional units.

**Supplementary Table S5: Relations between improvement in positive symptoms after treatment and striatal DS, GABA and glutamate levels**

| | $k_3$ | | GABA ACC | | $k_3$*Neuro-transmitter[a] | | Sex | | Glutamate thalamus | | Glutamate ACC | | Adjusted R-squared (%) |
|---|---|---|---|---|---|---|---|---|---|---|---|---|---|
| | **Coef.** | **p-value** | **Coef.** | **p-value** | **Coef.** | **p-value** | **Coef.** | **p-value** | **Coef.** | **p-value** | **Coef.** | **p-value** | |
| **Model 6**** <br> ΔPANSS pos = $k_3$ + GABA + GABA* $k_3$ + sex | -1.65 | **0.02** | 0.07 | 0.91 | -1.30 | **0.02** | -1.08 | 0.11 | - | - | - | - | 43.6% |
| **Model 7*** <br> ΔPANSS pos = $k_3$ + glu thal + glu thal* $k_3$ + sex | -2.03 | **0.008** | - | - | -0.10 | 0.86 | 1.82 | **0.015** | 0.50 | 0.45 | - | - | 24.9% |
| **Model 8*** <br> ΔPANSS pos = $k_3$ + glu ACC + glu ACC* $k_3$ + sex | -1.72 | **0.015** | - | - | -0.57 | 0.48 | -1.47 | 0.06 | - | - | 0.27 | 0.66 | 23.9% |

Supplementary Table S5 shows multiple linear regression models for the relation between symptoms improvement in positive symptoms after six weeks of treatment with a partial dopamine agonist (ΔPANSS positive) and dopamine synthesis in Nucleus Accumbens ($k_3$), GABA levels in anterior cingulate cortex (ACC), glutamate levels in ACC, as well as glutamate levels in thalamus in initially antipsychotic-naïve first-episode patients with psychosis. The dependent and independent variables are outlined for the separate models (without the intercept and β-coefficients due to space limitations). Sex was included as a covariate of no interest. * and ** for Model 6, 7, and 8 indicates that the overall models were statistically significant, and significant p-values for the independent variables are highlighted with bold.

[a]$k_3$*GABA for model 6, $k_3$*glu thal for model 7, and $k_3$*glu ACC for model 8.

*: $p<0.05$ and **: $p<0.01$.

Abbreviations: $k_3$: dopamine synthesis in Nucleus Accumbens; ACC: anterior cingulate cortex; PANSS: Positive and negative syndrome scale; Pos: positive; thal: left thalamus.
